# Quantitative definition of fever needs a change

**DOI:** 10.1101/2021.06.13.21258846

**Authors:** Nitin Kumar, Mayank Kapoor, Prasan Kumar Panda, Yogesh Singh, Ajeet Singh Bhadoriya

## Abstract

**Background:** The age-old definition of fever was derived using cross-sectional population surveying utilizing old techniques without considering symptomatology. However, the diagnosis of fever must be made only in the presence of associated symptoms that can distinguish it from the mere asymptomatic physiologic rise of temperature. Association of the temperature values with the symptoms to define the cut-off for fever is need of the hour.

**Methods:** A longitudinal study on the healthy population of Northen-India were followed-up over 1-year. Participants were advised for self-monitoring of oral temperature with a standard digital thermometer in either left or right sublingual pocket and record it in the thermometry diary. The study was considered complete if the participant had all the three phases of the study (i.e. non-febrile, febrile, and post-febrile phases) or completed the duration of the study.

**Results:** Per protocol analysis done for febrile participants (n=144, temperature recordings= 23851). The mean febrile phase temperature was 100.25 ± 1.44^0^F. A temperature of 99.1^0^F had maximum diagnostic accuracy for feeling feverish (98.2%), along with one (98.3%) or two (99%) associated symptoms. Summer and spring months showed higher temperatures (100.38 ± 1.44 v/s 99.80 ± 1.49, P<0.001), whereas no significant temperature difference could be noted amongst the gender.

**Conclusions:** A revised cut-off for the temperature to decide fever is hereby proposed: 99.10F along with one or two associated symptoms. This is going to redefine fever in the modern era completely.

## Introduction

“Humanity has but three great enemies: fever, famine, and war, and of these by far the greatest, by far the most terrible, is fever.” This statement by William Osler describes the paramount importance of fever since ancient medicine. The temperature has been one of the most important vital signs and recordings, it has been a critical component of good patient management. The core human body temperature depends on the appropriate functioning of the body.(1) Maintaining it within an optimal range is necessary for human life. It undergoes a regular circadian fluctuation of 0.5–0.7 °C, with the lowest in the early morning and highest in the evening. Similar temperature variation is also seen in the females during their menstrual cycle. The temperature may rise 0.6°C or more through the menstrual cycle.(2) Furthermore, the balance between heat production and heat loss determines the body temperature. Once this balance is lost, the temperature is raised in the body, known as fever. Hence, technically fever is a sign of some underlying pathology.

Wunderlich (1868) had defined the normal body temperature as 37°C (98.6°F). However, his methods were outdated. Mackowiak, Wasserman, and Levine (1992) set out to question this time-honored Wunderlich’s dictum. They did their cross-sectional study on young adults (younger than 40 years) using a standardized thermometer.(3) They concluded that 36.8°C (98.2°F) rather than 37.0°C (98.6°F) was the mean oral temperature of their participants; 37.7°C (99.9°F) rather than 38.0°C (100.4°F) was the upper limit of the normal temperature range. This approach to defining fever using cross-sectional temperature values of the population is incorrect. Temperature variation can be asymptomatic too, as described above in the case of females. Hence, the ideal way to define fever is to follow up healthy subjects prospectively, record their pre-febrile temperatures, the temperature during the time they feel feverish with other associated symptoms, and then measure the post-febrile phase temperatures.

Protsiv et al. (2020) hypothesized that the normal oral temperature of adults is lower than the established 37°C of the 19th century and concluded that body temperature has decreased over time in the USA using measurements.[4] Recent studies suggest that normal temperature has invariably decreased by 0.03°C per birth decade probably due to lowered metabolic rate and infections, henceforth drifting down the normal morning body temperature to less than <98.6°F over the last two centuries (4–8). The influence of age, time of day, gender, and economic development preclude an updated definition of fever.

Thereby, we did a longitudinal study on healthy participants using a standardized electronic thermometer in the left or right posterior sublingual pocket and analyzed the associated symptomatology to derive a new symptom-associated definition of fever.

## Methods

### Study setting, design, and participants

The study was conducted at All India Institute of Medical Sciences Rishikesh (AIIMS), a tertiary healthcare center in the state of Uttarakhand, India. It was a longitudinal study conducted over 15 months from July 2019 to September 2020.

Lists of employees and students were obtained from the human resource department and registrar’s office. Information of participating family members was obtained from consenting employees and students and was selected via a simple random sampling method using the computer. If the participant did not give consent for the study then the next person on the list was selected. We had taken the standard deviation according to a study done by Mackowiak et al as 0.7 and employing T-distribution to estimate sample size, the study would require a sample size of 192 with 95% confidence and a precision of 0.1 (3,09).

The participants were recruited based on the following inclusion and exclusion criteria after taking informed consent.

- Inclusion criteria: healthy staff and students of AIIMS and their accompanying family members between 4-100 years and who agreed to work/study at AIIMS during the duration of the study (1year).
- Exclusion criteria: any individuals with any diagnosed or suspected disease (any acute infectious or non-infectious illness (including trauma) within last 1month and post-partum period up to 8 weeks; any known case of or past history of chronic illness - infective (e.g. tuberculosis, kala-azar, brucellosis, infective endocarditis, HIV/AIDS, hepatitis B/C/D etc.), rheumatological (e.g. RA, SLE, vasculitis etc.), chronic liver disease, chronic kidney disease, cardiovascular disease (e.g. systemic hypertension, coronary artery disease, valvular heart disease, pulmonary arterial hypertension, peripheral vascular disease etc.), chronic lung disease (e.g. any obstructive or restrictive airway diseases), endocrinopathy (e.g. diabetes mellitus, diabetes insipidus, hypo/hyperthyroidism etc.), gastro-intestinal disease (e.g. dyspepsia, inflammatory bowel disease, malabsorption syndromes etc.), neurologic disorders (e.g. epilepsy, stroke, dementia, movement disorder, degenerative disorder, cerebral palsy etc.), psychiatric disorder (e.g. mood disorders, psychosis, dependence syndrome(s) etc.), dermatological diseases (e.g. bullous disorders, psoriasis, tinea etc.), any malignancy (treated or otherwise), recent history of vaccination in last 6 months, and ankyloglossia.

### Interventions

Detailed clinical evaluations (history and examination) were done. Basic investigations (with the last 1year; as per institute recruitment policy): ECG, chest X-ray, viral markers (anti-HIV-1 and 2, HBsAg, anti-HCV), urine routine, complete blood count, fasting blood glucose, liver and kidney function tests were collected from medical record section after approval from Institute Ethical Committee, All India Institute of Medical Sciences (AIIMS), Rishikesh (No. 235/IEC/PGM/2019). If any abnormality was detected, they were excluded without sharing details.

The study was done in three phases: first phase (non-febrile phase), second phase (febrile phase), and third phase (post-febrile phase). The subjective sensation of feeling feverish by the participant was taken to define the febrile phase’s onset, if it persisted for >24 hours or the participant had to take antipyretics. One clinical thermometry diary, a ball pen, a and standard electronic thermometer (Dr.Morepen Digiflexi Flexi Tip Thermometer (MT222)) were provided to all participants. The detailed procedure was explained the and first reading was verified and they were followed up fortnightly physically, and reminded weekly through WhatsApp and telephonically. Procedure to be followed while measuring temperature was explained as follows:

- Wash your hands with soap and water.
- Use a clean thermometer, one that has been washed in cold water, cleaned with rubbing alcohol, and then rinsed to remove the alcohol.
- Please ensure that no physical exertion in the past 30min, avoid any cold or hot beverage or food in the past 30min, no smoking during the past 30 min.
- Sit in a cool and calm environment for at least 30 min.
- Keep the thermometer in an oral left or right posterior sublingual pocket and keep your mouth closed during this time.
- Hold the thermometer in the same spot until it makes a beeping noise when the final reading is done.
- Readings will continue to increase and the °F (& °C) symbol will flash during measurement.
- Kindly record the temperature on the Fahrenheit scale and the time in the thermometry diary.
- Rinse the thermometer in cold water, clean it with alcohol, and rinse again and store it in a safe place for the next readings.
- Three readings were taken once after waking up (AM), once in the afternoon (AN; 12-3 PM), and once before sleeping (PM), also requested to mark any symptoms from the checklist simultaneously.
- There were 3 days of more frequent temperature charting (every 2^nd^ hour, except sleeping time) during the non-febrile phase and 2 days of frequent temperature charting during post febrile phase, and for all the days during the febrile phase
- For family members having children and old age persons, staff or students measured the temperature and recorded.

Self-recording of data was done in the provided clinical thermometry diary and the same was assessed fortnightly by the investigator(s) for troubleshooting and to see the status of recording. Diary was collected once the participant had all the three phases of the study or at the end of the study if no febrile episodes.

### Comparator/outcomes

There was no comparator except among three phases of temperature recordings.

Participants were further divided into 4 subgroups based on seasonal months: November-January (represented coldest months of the year); February-April (representing spring months); May-July (representing hottest months); August-October (representing autumn months). A maximum of 45 days of data was taken immediately before the febrile phase, during the non-febrile phase of per-protocol analysis. A maximum of 10 days of data was taken immediately after the febrile phase during the post-febrile phase, and complete data of the febrile phase was taken for analysis. The whole of the frequent temperature readings (i.e. two hourly temperature records) was taken for analysis for all the three phases especially to see variations.

### Statistical analysis

The data was entered in the excel sheet and primary outcomes were analyzed as per-protocol analysis (for those participants who had all the three phases in the study) using Statistical Package for Social Sciences (SPSS) Version 23. Categorical variables were presented as number and percentage (%) and continuous variables as mean (SD). The Kolmogorov-Smirnov test tested the normality of data, and if rejected, a non-parametric test was used. Quantitative variables were compared using independent t-test/Wilcoxon–Mann–Whitney test (when the data sets were not normally distributed) between two groups and Kruskal Wallis test between three and more groups. The continuous variables, those were not normally distributed, analyzed using Shapiro-Wilk Test. To define fever cut-offs with respect to symptoms, receiver operating characteristic (ROC) curve analysis was done, and the cut-off was taken as the point with maximum diagnostic accuracy. Taking confidence level as 95%, a P-value <0.05 was taken as statistically significant.

## Results

Three hundred fifty (350) participants were screened, and a total of 250 participants consented to be a part of the study. One hundred forty-four (144) were included in the per-protocol analysis (Figure 1: Study flow). The participants included healthy subjects with a mean age of 24.24 ± 5.92 years (8-58 years). 26% (56) were < 20 years, 72.1% (155) belonged to the age group 20-40 years, and 1.9% (4) were >40 years of age. 52.1% (112) were males. The associated febrile symptoms were observed (Table 1).

**Table 1:** Associated symptoms and their frequency in the cohort.

| Symptoms | Frequency (%) |
| --- | --- |
| Chills/Shivering | 669 (44.6%) |
| Dehydration/Increased Thirst | 72 (4.8%) |
| Dizziness | 81 (5.4%) |
| Fatigue | 755 (50.3%) |
| Feeling Malaise/General Weakness | 700 (46.7%) |
| Headache/Head Heaviness | 705 (47.0%) |
| Increased Breathing Rate | 556 (37.1%) |
| Increased Sweating | 645 (43.0%) |
| Irritability | 583 (38.9%) |
| Loss Of Appetite | 698 (46.5%) |
| Muscle Cramps/Muscle-aches | 684 (45.6%) |
| Nausea | 638 (42.5%) |
| Restlessness/Anxiety/Palpitations | 548 (36.5%) |
| Warmth | 709 (47.3%) |

**Figure 1:**
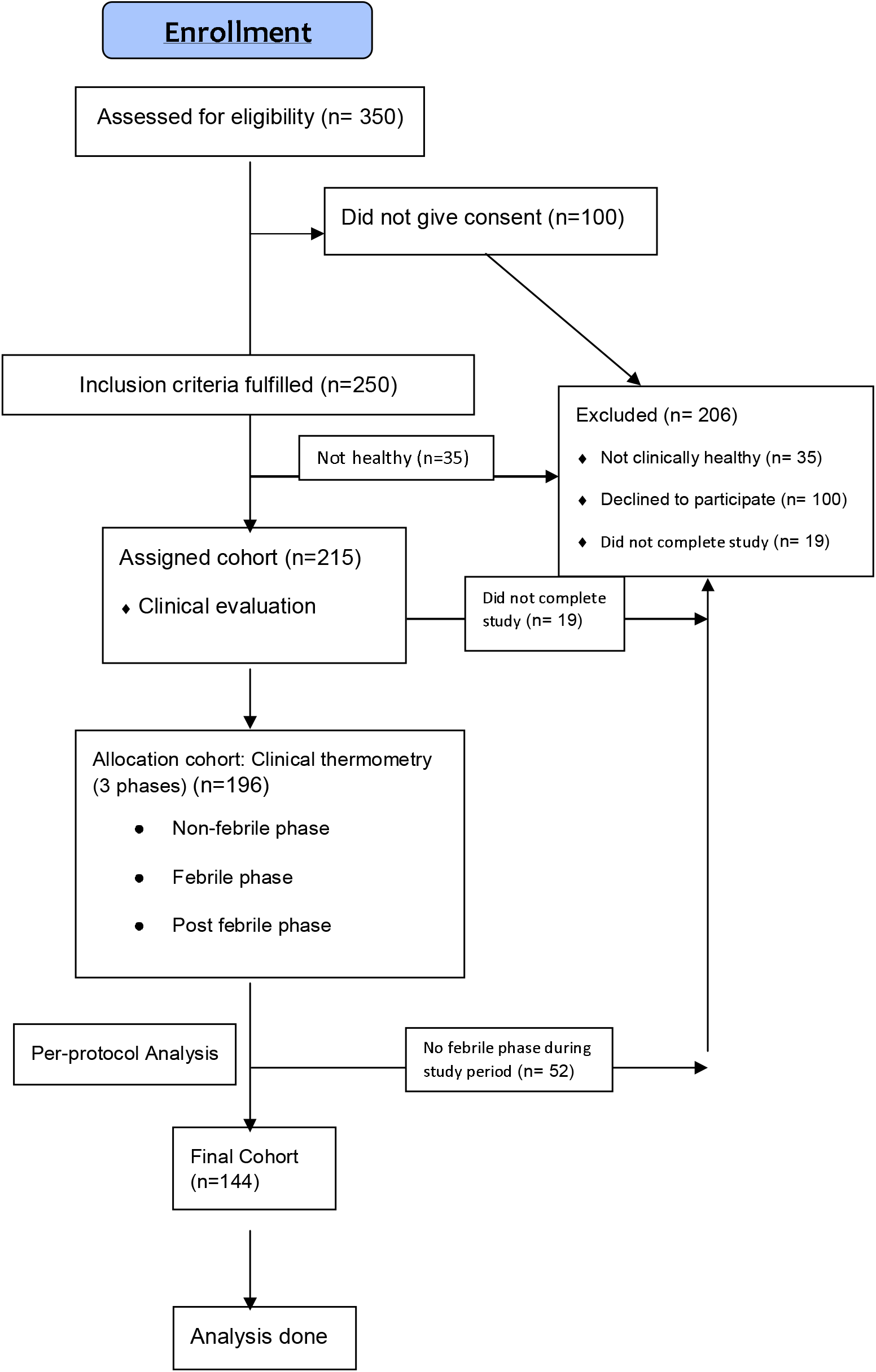
The study flow

### Definition of fever and associated symptoms

The temperature cut-offs for feeling feverish were determined based on ROC analysis with diurnal, seasonal, and gender variations (Figure 2 A/B/C, Table 2).

**Table 2:** Sensitivity, specificity, and diagnostic accuracy of various temperature cut-offs for determination of fever.

| for determination of fever |  |  |  |  |
| --- | --- | --- | --- | --- |
| Group | Temperature Cut-Off (°F) | Sensitivity | Specificity | Diagnostic accuracy |
| Feeling feverish | 98.5 | 91% | 75% | 75.4% |
|  | 98.8 | 89% | 88% | 88% |
|  | **99.1** | **83 %** | **99 %** | **98.2%** |
|  | 99.3 | 79% | 99% | 97.6% |
|  | 99.5 | 78% | 99% | 97.5% |
| Feeling feverish and one more associated symptom | 98.5 | 94.7% | 70% | 70% |
|  | 98.8 | 93% | 84% | 84% |
|  | **99.1** | **86 %** | **99 %** | **98.3%** |
|  | 99.3 | 82% | 98.5% | 97% |
|  | 99.5 | 81% | 98.8% | 97.8% |
| Feeling feverish and two more associated symptoms | 98.5 | 99.9% | 70% | 71% |
|  | 98.8 | 96% | 84% | 84% |
|  | **99.1** | **88 %** | **99 %** | **99%** |
|  | 99.3 | 87% | 99% | 98.9% |
|  | 99.5 | 85% | 99% | 98.8% |

**Table 3:**
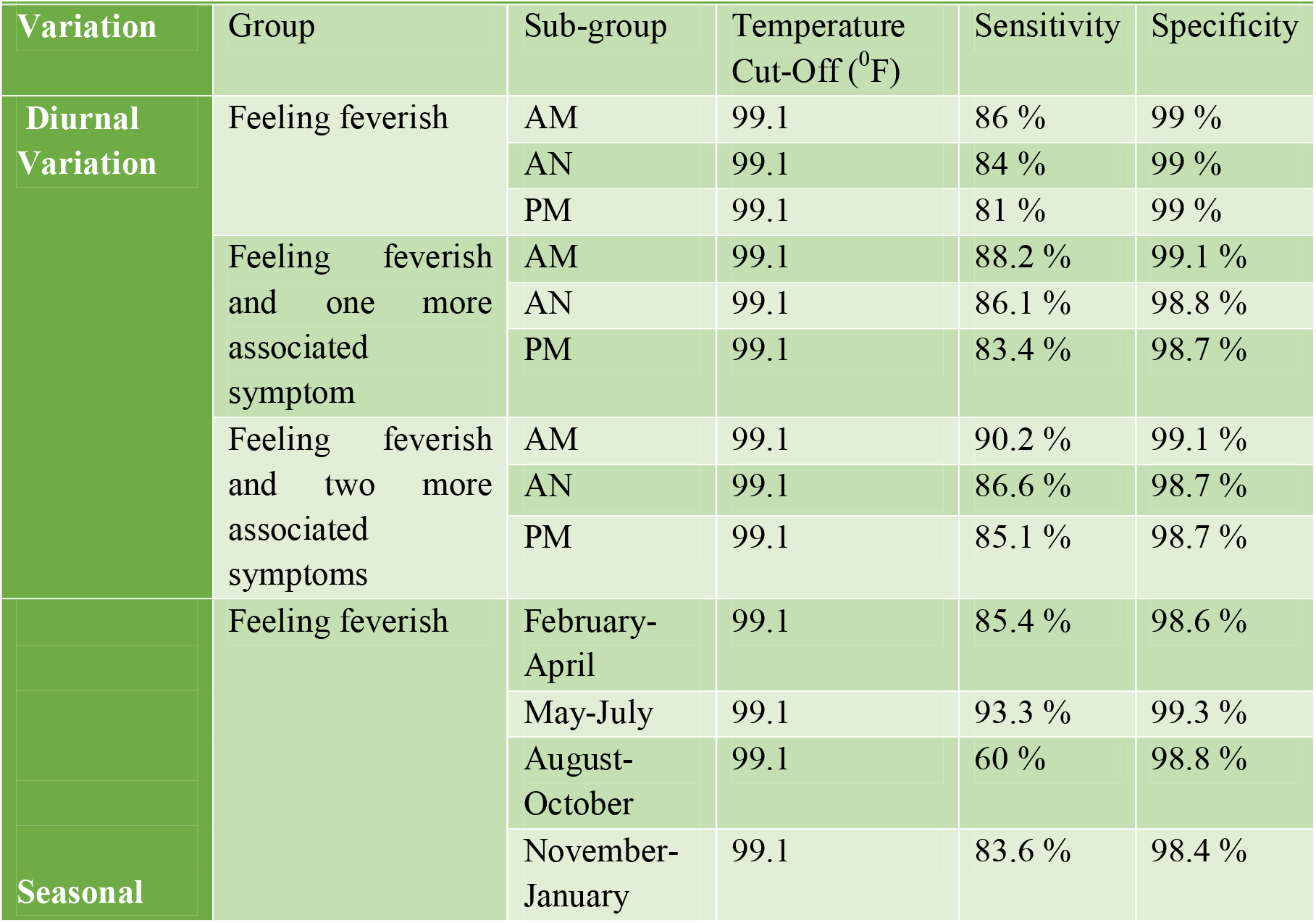
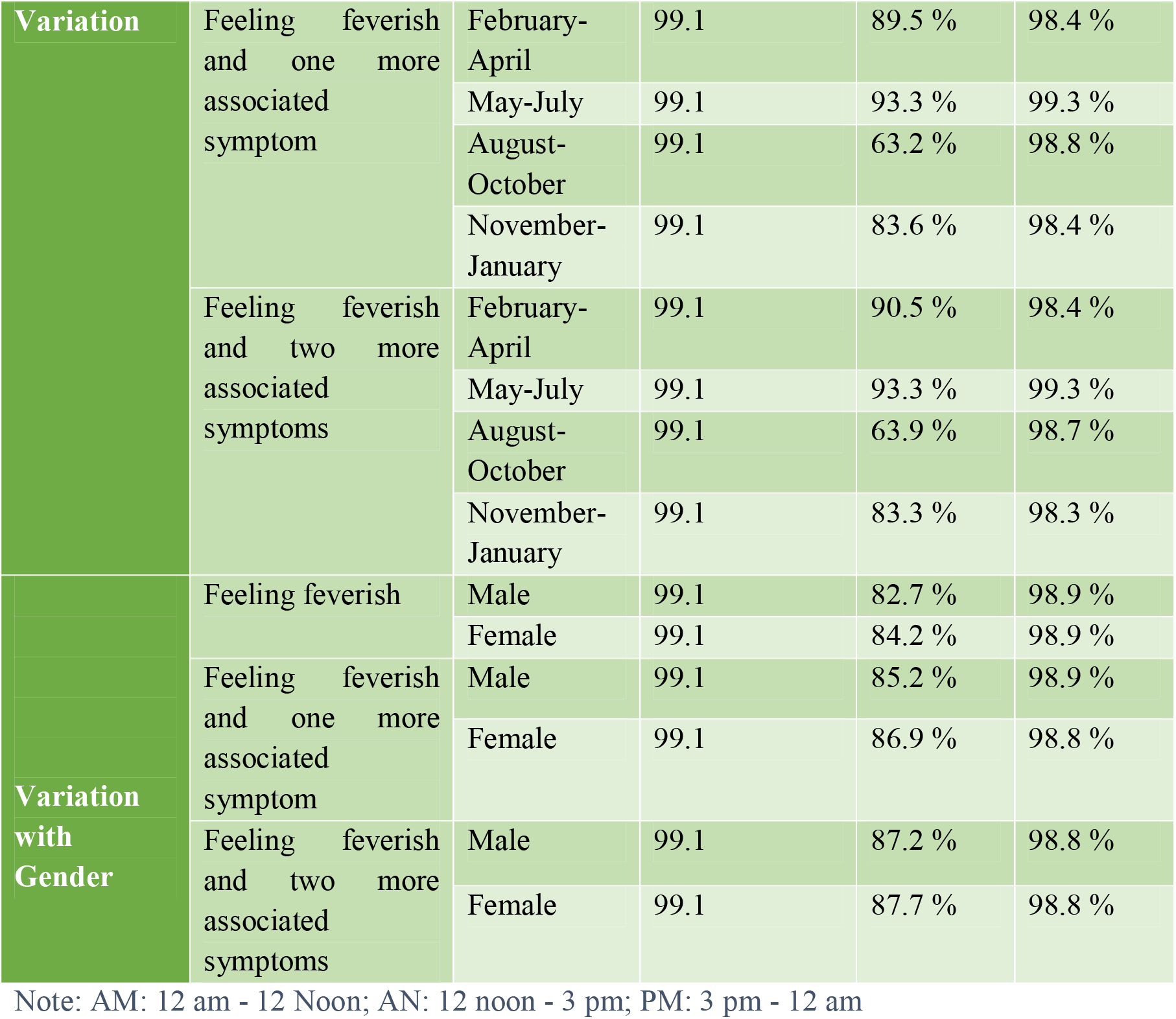
Sensitivity and specificity of temperature cut-off 99.1 ^0^F in determining fever amongst subgroups.

**Figure 2:**
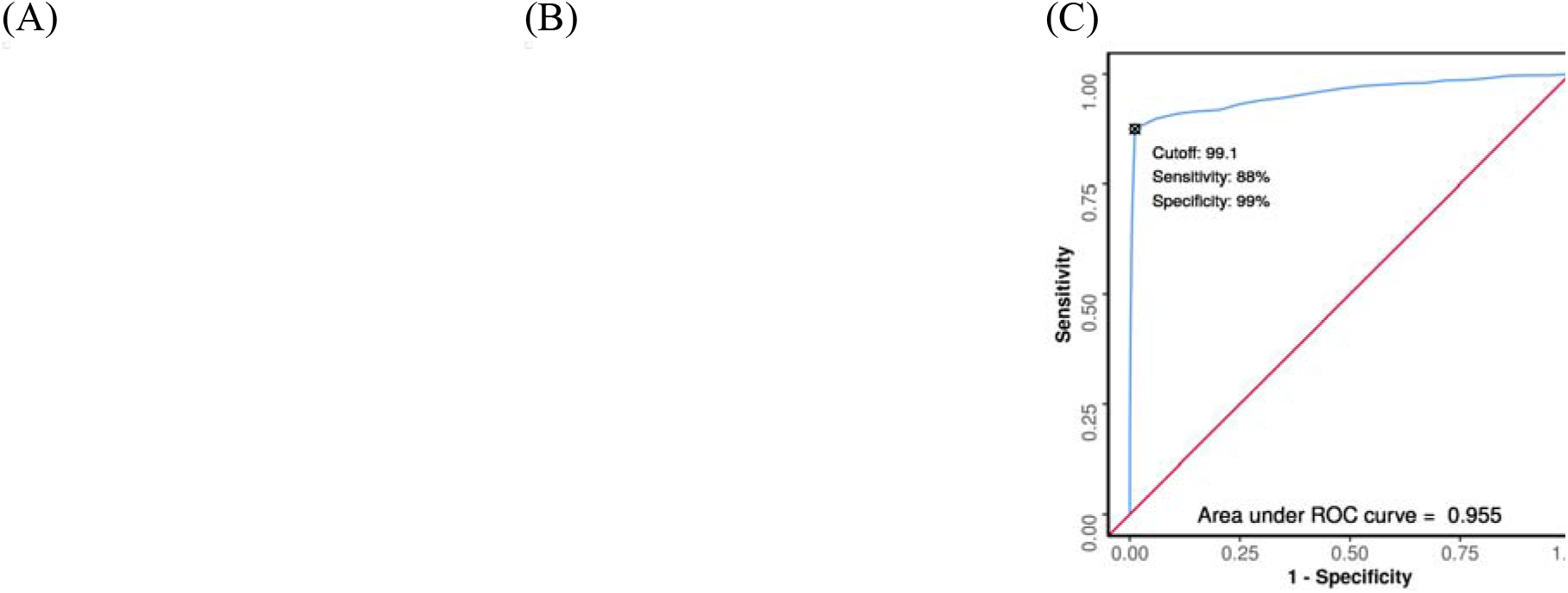
ROC Curve Analysis showing diagnostic performance of (A) Temperature in Predicting Feeling Feverish; (B) Temperature + 1 More Symptom in Predicting Feeling Feverish; (C) Temperature + 2 More Symptoms in Predicting Feeling Feverish

## Discussion

We analyzed 6544 temperature readings of the 144 healthy participants longitudinally over 1-year. This longitudinal study of a healthy population, mainly in the adult age group, demonstrates that a temperature of 99.1^0^F has the highest diagnostic accuracy in predicting fever (98.2%), which increases further when associated with one (98.3%) or two (99%) additional symptoms. The diagnostic accuracy of temperature measurement and the associated symptomatology for fever prediction is highest in the morning compared to the afternoon or evening. We also found that the predictive ability was maximum in the summer months (May-July) compared with spring, winter, and autumn, respectively. Our criteria demonstrated higher sensitivity amongst the females than the males. Accuracy increased with the increase in the number of associated symptoms.

An AM temperature of >37.2°C (>98.9°F) or a PM temperature of >37.7°C (>99.9°F) defines fever, and the American College of Critical Care Medicine, the International Statistical Classification of Diseases, and the Infectious Diseases Society of America define fever as a core temperature of 38.3°C (100.9°F) or higher, just above the upper limit of a normal human temperature, irrespective of the cause.(10,11) Our quantitative diagnostic study considers the associated symptomatology to determine the temperature cut-off for fever and is the first to be reported. As mentioned before, all previous studies on the definition of fever were cross-sectional, and no study took into account the symptomatology along with the quantification of fever. Hence, we believe this is a landmark study that defines fever accordingly when the person also has the associated symptoms of fever as mere temperature rise can be physiological also.

Usually, the body temperature rises as the day passes.(12) This formed the basis of the old fever definition having a lower threshold for the morning temperature than the evening. However, our study demonstrated a reversal in this pattern, with AM temperature 100.39 ± 1.44°F, AN 100.11 ± 1.52°F, and PM 100.06 ± 1.44°F (P<0.001). This can be explained since patients were allowed to take antipyretics, which most prefer to take during the day. This led to a lower value of the evening temperatures in the febrile phase, forming a limitation of our study.

Renbourn and Bonsall (1946) in British India found out that oral temperatures higher than those accepted as usual for temperate climates were not uncommon during the summer in North India. Oral and rectal temperatures of both Indian and British troops during the summer months in North India showed levels above those accepted as normal for temperate climates.(13) This again demonstrates that the temperature per se is just a quantitative variable that can also undergo fluctuations with the outside seasonal variation, further strengthening our study’s importance. A mere rise of temperature value should not be called fever, but this should be termed as fever when combined with the associated symptoms. In our study also, the temperature values were highest in the spring (100.38 ± 1.44°F) and summer (100.26 ± 1.40°F) months as compared to winter (100.13 ± 1.42°F) and autumn (99.80 ± 1.49°F) (P<0.001). Females are considered to have higher baseline temperature, although we could not find any significant temperature difference between the two sexes in our study.

The study has limitations also. The sample was unicentric, difficult for generalization of results, thus a larger multi-centric study is required. The vulnerable group of the population, elderly and children, could not be included in the study desirably. The participants were defined healthy based on history and pre-defined biochemical and laboratory parameters, therefore, indolent chronic infections and sub-clinical non-infectious illnesses couldn’t be ruled out. No physical way of checking the adherence to the advised procedure for the temperature measurement was there. The participants were reviewed followed up fortnightly. So, strategies to measure directly observed temperature may require. Compliance was overall poor (∼38%). The oral temperature in the left or right sub-lingual pocket is close representative of core body temperature, but not exactly the core body temperature. The ongoing COVID-19 pandemic might have influenced the results. The participants constituted a high-risk populace for infectious agents and stress. As mentioned, patients were allowed to take antipyretics; hence the temperature values could be lower with drug use. Another major limitation is that the febrile phase’s categorization in our study was solely based on the subject’s subjective sensation of feeling feverish. As it is a subjective sensation, it can vary from person to person. Nevertheless, this in itself forms the basis of our study that fever is a sign that varies from person to person, and it is not merely a numerical cut-off that can be generalized to the whole population.

## Conclusion

We propose an oral temperature cut-off of 99.10F, along with one or more associated symptoms, to accurately predict fever with a sensitivity of 88% and specificity of 99%. This finding completely changes our understanding of fever and calls for a universal change in the definition of the same.

## Data Availability

It will be available with corresponding author.

## Contributors

NK contributed to the data collection, data analysis, and was involved in manuscript writing. MK, YS, and ASB contributed to the data analysis and were involved in reviewing the draft. PKP gave the concept, interpreted analysis, critically reviewed the draft, and approved it for publication along with all authors.

## Data sharing

It will be made available to others as required upon requesting the corresponding author.

## Acknowledgment

None

